# Long-term retention and associated factors among volunteer community health workers in southwestern Uganda: a 17-to-21-year retrospective cohort study

**DOI:** 10.64898/2026.10.20.26365976

**Authors:** Edison Byamugisha, Hamson Kanyesigye, Teddy Kyomuhangi, Glorious Atukunda, Jenn Brenner, Jerome Kabakyenga

**Affiliations:** Department of Maternal Newborn and Child Health Research, Mbarara University of Science and Technology, Mbarara, Uganda; Department of Community Health, Mbarara University of Science and Technology, Mbarara, Uganda; Department of Pediatrics and Community Health Sciences, Cumming School of Medicine, University of Calgary, Calgary, Canada; Department of Quality Assurance, Mayanja Memorial Medical Training Institute, Mbarara, Uganda

## Abstract

Community health workers (CHWs) are central to primary healthcare delivery in low-resource settings, yet evidence on what sustains their engagement beyond a decade is limited, particularly in sub-Saharan Africa. This study examined long-term retention and its associated factors among volunteer CHWs in southwestern Uganda, 17 to 21 years after their initial selection and training. We conducted a quantitative retrospective cohort study using a census approach in April 2024 across three districts in south western Uganda where a cohort of CHWs were selected and trained between 2004 and 2008. Data were collected in April 2024 using a pretested, structured, interviewer-administered questionnaire. Data were analysed using STATA version 14.1. Descriptive statistics characterized the study population, stratified by retention status. Bivariate and Multivariable Poisson regression models were used to identify factors independently associated with long term retention of CHWs.

From the 404 CHWs selected and trained in the period 2004-2008, 265 (65.6%) were still resident in the study districts. Of 265 CHWs, 161 (60.8%) were categorized as retained. Factors independently associated with retention were: peer support (adjusted incidence rate ratio [IRR] = 2.21, 95% confidence interval [CI]: 1.48–3.32, p < 0.001), always having access to reporting tools (adjusted IRR = 1.46, 95% CI: 1.10–1.95, p = 0.01), and intermittent access to reporting tools (adjusted IRR = 1.39, 95% CI: 1.08–1.79, p = 0.01). The most common reasons for exit were family duties, poor personal health, and community rejection. Over 60% of trained CHWs remained active after nearly two decades, indicating that volunteer CHW programmes can achieve sustained long-term engagement. Peer support and reliable access to reporting tools were the strongest modifiable predictors of retention. Strengthening these low-cost programmatic supports may be a cost-effective strategy for improving CHW retention in similar low-resource settings, and programme managers should prioritise structured peer-support networks and a consistent supply of reporting tools.

## Introduction

Improving maternal, newborn and child health (MNCH) and survival remains a global priority(1). Community health workers (CHWs) extend health promotion, referral and curative support to underserved and remote populations, and a substantial evidence base supports their contribution to reducing child morbidity and mortality in sub-Saharan Africa (2). This evidence has driven widespread scale-up of CHW programmes across low- and middle-income countries (LMICs)(3), where CHWs are now embedded within national health system strategies and viewed as central to progress towards achieving the Sustainable Development Goals(4).

However, volunteer CHW programmes face a persistent sustainability challenge: retention (5). Attrition/exit undermines programme continuity, erodes community trust built through long-standing relationships, and increases the cost of continual replacement and retraining(6). Most published evidence on CHW retention covers the short to medium term - typically one to five years - with far fewer studies extending beyond a decade, particularly in sub-Saharan Africa(5). In Uganda, the Ministry of Health established the national Village Health Team strategy in 2001 to formalize community-level volunteer engagement. In southwestern Uganda, the Healthy Child Uganda (HCU) partnership - between Mbarara University of Science and Technology and Canadian academic partners - piloted a volunteer CHW model focused on MNCH promotion beginning in 2004, selecting and training 404 CHWs across three districts by 2008(7).More information about this partnership is available at www.healthychilduganda.org/resources. An early impact evaluation of this pilot cohort found the model to be associated with reduced under-five morbidity and reported encouraging CHW retention over the initial years of implementation (7). A subsequent evaluation of this same cohort reported very high retention -91% at 2 years and 86% at 5 years and identified non-financial incentives, particularly peer and team support, as more salient to CHW motivation than monetary compensation(8). A later scale-up of the HCU model to a much larger cohort (n = 2317) similarly reported high medium-term retention (84% at 5 years), with sex and age, rather than financial or workload factors, as the main correlates of exit(9). Complementary qualitative work within the same programme linked supportive supervision and constructive peer relationships to sustained CHW performance(10).

Despite this substantial body of evidence from the HCU programme and elsewhere on short- to medium-term CHW retention, no study has followed a volunteer CHW cohort in this setting beyond one decade to determine whether, and why, engagement persists over the very long term. This is an important gap: whether early predictors of retention, such as peer support and non-financial incentives, remain relevant fifteen to twenty years after training has direct implications for the design of sustainable, low-cost CHW programmes intended to operate over the long term. This study addresses this gap by revisiting the original 2004–2008 HCU pilot cohort 17–21 years after training, to determine the proportion still actively engaged and to identify the programmatic factors independently associated with this long-term retention. This study found that 60.8% of the of the 265 CHWs selected and trained in 2004 to 20028 but still resident in the 3 districts were categorized as retained in their role 17-21 years later.

## Materials and methods

### Study design and setting

This was a quantitative retrospective cohort study using a census approach, consistent with the Strengthening the Reporting of Observational Studies in Epidemiology (STROBE) guideline for cohort studies (11). The study setting was three districts of southwestern Uganda - Mbarara, Rubirizi and Rwampara - where Healthy Child Uganda implemented its first volunteer CHW pilot programme between February 2004 and January 2008(7,8). These districts represent typical rural and peri-urban settings in southwestern Uganda, providing locally specific data on long-term retention across varied implementation contexts.

### Study population and eligibility

The target population was CHWs selected and trained by Healthy Child Uganda between 2004 and 2008 who were still residing in the three districts at the time of data collection (April 2024). A census approach was chosen because the total eligible population (n = 404) was manageable and allowed disaggregated analyses by district and subgroup. CHW coordinators from each parish assisted investigators in verifying and updating CHW registers maintained since 2004. Inclusion criteria were: CHWs trained by Healthy Child Uganda in 2004–2008, still residing in the study areas, and willing to provide informed consent.

### Data collection

Data were collected in 1^st^ April 2024 to 30^th^ April 2024 using a pretested, structured, interviewer-administered questionnaire with closed-ended items, developed in English and translated into the local language (Runyankole). The principal investigator (EB) conducted all interviews.

### Outcome definition

Retention was defined as regular submission of monthly activity reports in the two months immediately preceding data collection (April 2024)(9). This definition reflects field practice, whereby CHWs who did not submit monthly workload reports for two consecutive months are considered to have dropped out (9). Non-submission for two months was classified as attrition/exit.

### Statistical analysis

Data were analysed using STATA version 14.1 (StataCorp, College Station, TX). Descriptive statistics (frequencies, percentages, and medians with interquartile ranges [IQR]) characterised the study population, stratified by retention status. Bivariate Poisson regression with robust standard errors generated unadjusted incidence rate ratios (IRR) and 95% confidence intervals for each candidate predictor.

Multivariable model selection followed Hosmer and Lemeshow’s purposeful selection approach (12): variables with p < 0.20 in bivariate analysis, or known confounders from the literature, were entered into an initial multivariable model. Variables with p > 0.05 were removed sequentially, then re-introduced individually to evaluate potential confounding (defined as a >10% change in the IRR of any retained variable). An interaction term between community type and district was tested using a log-likelihood ratio test and found non-significant; it was dropped from the final model. Robust standard errors were retained to account for potential clustering effects.

### Ethical considerations

Ethical approval was obtained from the Research Ethics Committee of Mbarara University of Science and Technology (MUST-2023-1183) and the Uganda National Council for Science and Technology (SS2542ES). Community entry approvals were obtained from the Chief Administrative Officers of the three districts. Participants provided written informed consent before enrolment. Each participant was assigned a unique code; names were not recorded. Participants received a transport reimbursement of UGX 10,000 (USD 2.70).

## Results

### Cohort disposition

Of 404 CHWs originally trained between 2004 and 2008, 265 (65.6%) were still resident in the study areas and formed the study sample. Of these, 161 (60.8%) were classified as retained (active) and 104 (39.2%) as having exited the programme at the time of data collection (**see *Table 1***).

**Table 1.** Cohort disposition and overall retention rate.

| Cohort stage | N | % |
| --- | --- | --- |
| Still resident in study areas (study sample) | 265 | 100 |
| Active / Retained | 161 | 60.8 |
| Inactive / Exited | 104 | 39.2 |

*Retention was defined as regular submission of monthly reports in the two months preceding April 2024.*

### Characteristics of the participants

The majority of participants were female (72.1%, n = 191), married (84.2%, n = 223), and had primary education or less (57.4%, n = 152). The median age was 55 years (IQR: 50–60) and 90.6% (n = 240) lived in rural areas. Demographic characteristics were broadly similar between retained and exited CHWs, with no statistically significant differences observed for sex (IRR = 0.88, 95% CI: 0.70–1.11, p = 0.29), age (IRR = 1.01, 95% CI: 1.00–1.02, p = 0.22), or rural residence (IRR = 0.94, 95% CI: 0.69–1.29, p = 0.72).

Regarding programme support, 86.8% (n = 230) reported never receiving monetary allowances. Peer support was available to 72.5% (n = 192); 88.2% of retained CHWs had peer support compared with only 48.1% of those who exited (IRR = 2.84, 95% CI: 1.91–4.22, p < 0.001). Access to reporting tools also varied considerably between groups, with both consistent access (21.7% retained vs. 14.4% exited; IRR = 1.61, 95% CI: 1.22–2.11, p = 0.001) and intermittent access (46.6% retained vs. 22.1% exited; IRR = 1.76, 95% CI: 1.39–2.22, p < 0.001) significantly associated with retention. **Table 2** presents the distribution of key socio-demographic and programme-related characteristics by retention status, together with results of bivariate analyses.

**Table 2.**
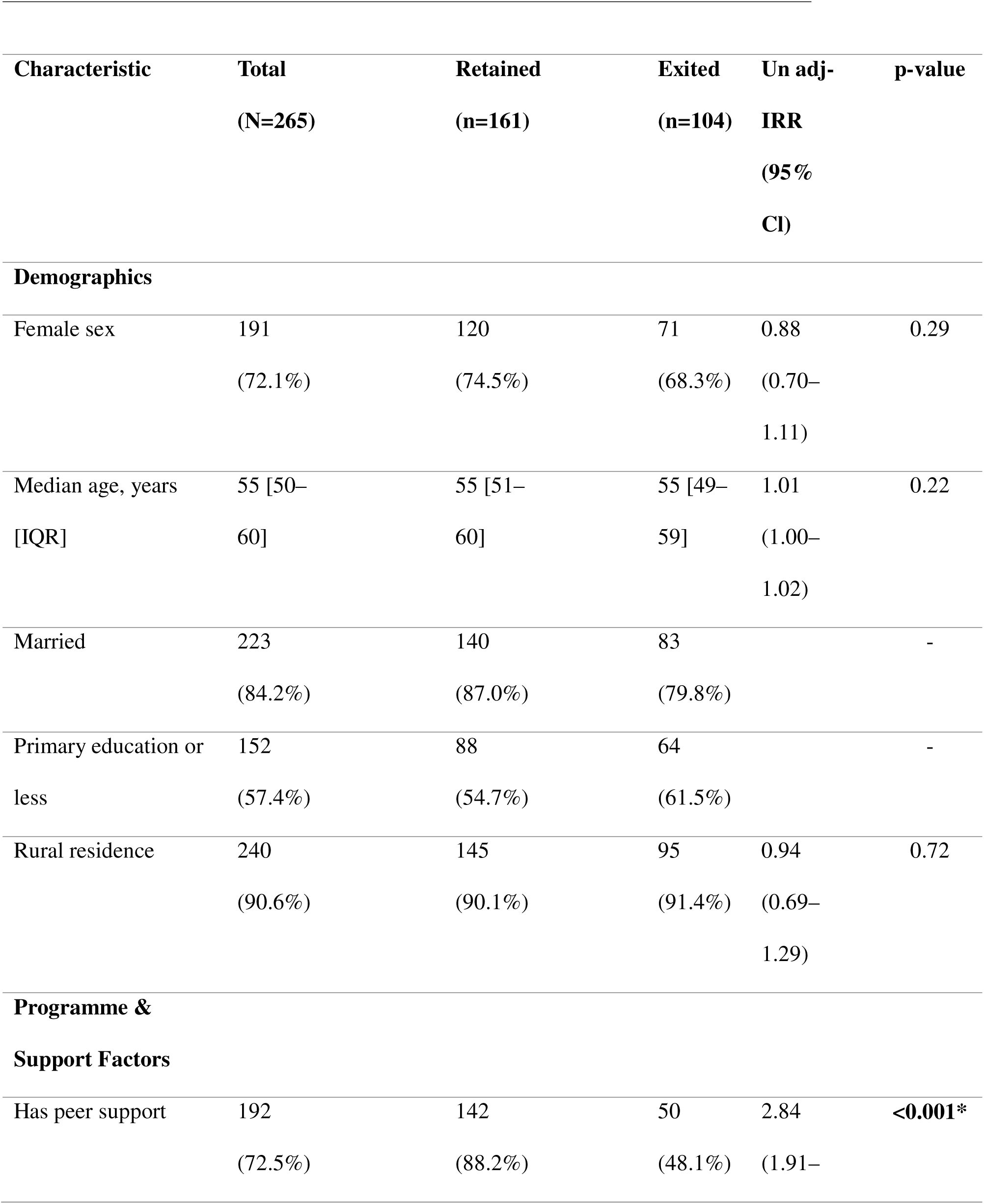
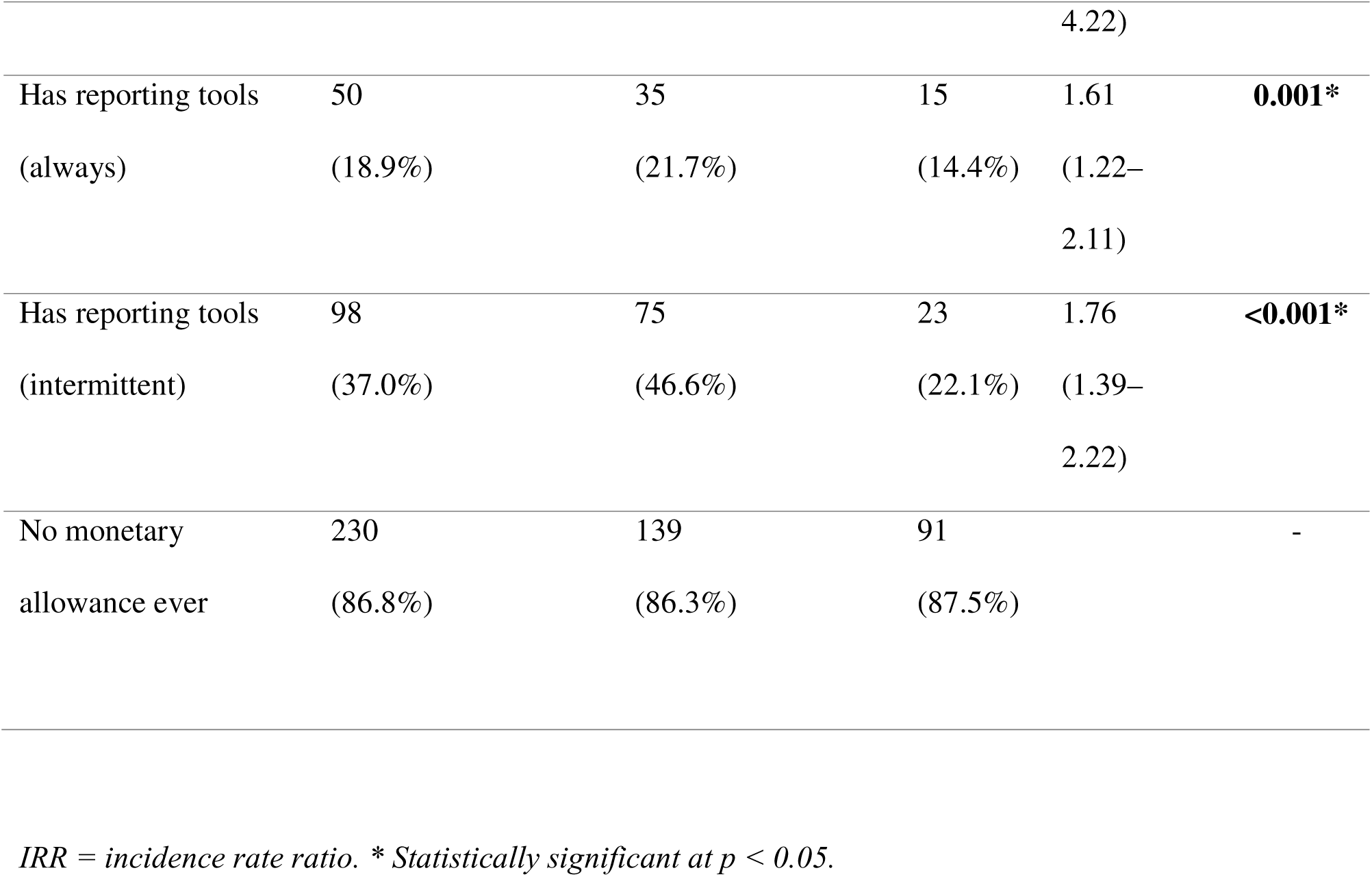
Socio-demographic and programme characteristics of CHWs by retention status.

**Table 3.**
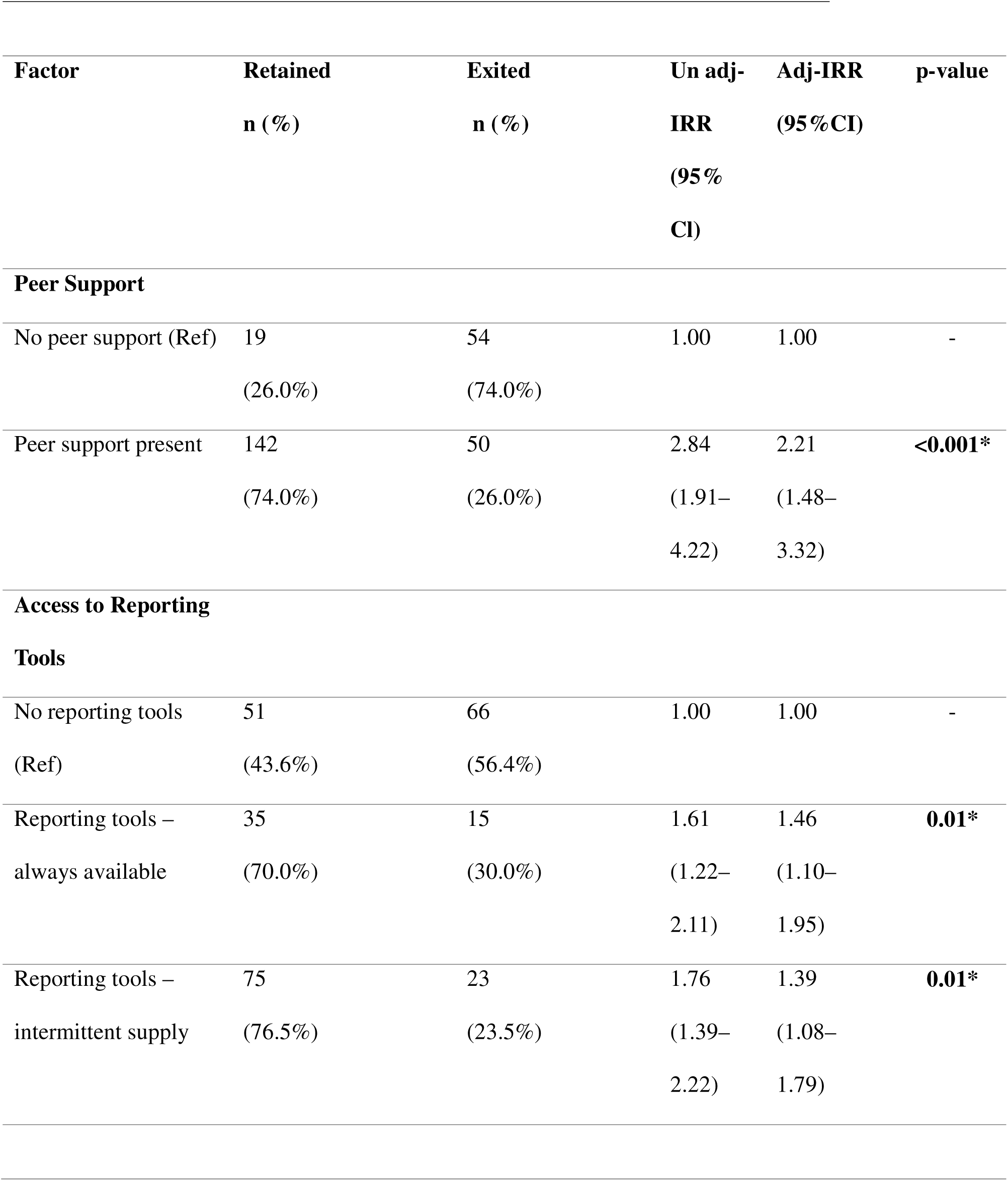
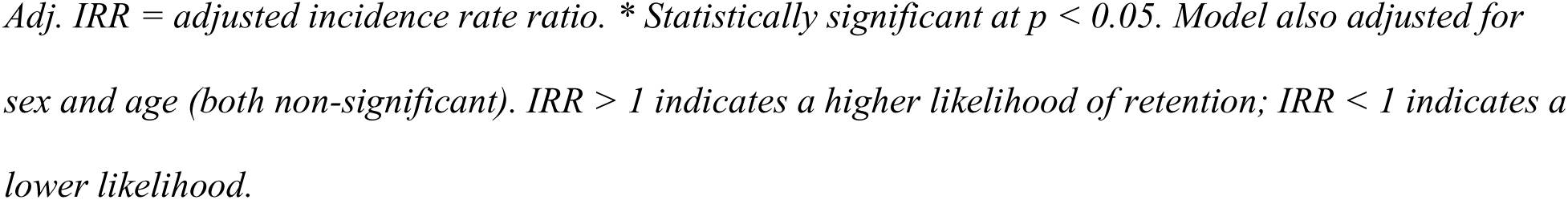
Multivariable analysis: factors independently associated with CHW retention (poission regression with robust standard errors n=265)

| **Factor** | **Retained**<br>**n (%)** | **Exited**<br>**n (%)** | **Un adj-**<br>**IRR**<br>**(95%**<br>**CI)** | **Adj-IRR**<br>**(95%CI)** | **p-value** |
| --- | --- | --- | --- | --- | --- |
| **Peer Support** |  |  |  |  |  |
| No peer support (Ref) | 19<br>(26.0%) | 54<br>(74.0%) | 1.00 | 1.00 | - |
| Peer support present | 142<br>(74.0%) | 50<br>(26.0%) | 2.84<br>(1.91–<br>4.22) | 2.21<br>(1.48–<br>3.32) | **<0.001\*** |
| **Access to Reporting**<br>**Tools** |  |  |  |  |  |
| No reporting tools<br>(Ref) | 51<br>(43.6%) | 66<br>(56.4%) | 1.00 | 1.00 | - |
| Reporting tools –<br>always available | 35<br>(70.0%) | 15<br>(30.0%) | 1.61<br>(1.22–<br>2.11) | 1.46<br>(1.10–<br>1.95) | **0.01\*** |
| Reporting tools –<br>intermittent supply | 75<br>(76.5%) | 23<br>(23.5%) | 1.76<br>(1.39–<br>2.22) | 1.39<br>(1.08–<br>1.79) | **0.01\*** |

Adj. IRR = adjusted incidence rate ratio. \* Statistically significant at $p < 0.05$ . Model also adjusted for sex and age (both non-significant). $IRR > 1$ indicates a higher likelihood of retention; $IRR < 1$ indicates a lower likelihood.

### Multivariable analysis of factors associated with retention

After adjusting for potential confounders, two modifiable factors remained independently and significantly associated with retention (**see *Table 2***). Peer support was the strongest predictor of retention: CHWs with access to peer support were more than twice as likely to be retained compared with those without (adjusted IRR = 2.21, 95% CI: 1.48–3.32, p < 0.001). Access to reporting tools was also a significant predictor: CHWs with consistent access were 46% more likely to be retained compared with those without access (adjusted IRR = 1.46, 95% CI: 1.10– 1.95, p = 0.01), while those with intermittent access were 39% more likely to be retained (adjusted IRR = 1.39, 95% CI: 1.08–1.79, p = 0.01).

### Reasons for exit

Among the 104 CHWs who exited, reasons were broadly classified as role-related or role-unrelated/life-circumstance (**see *Table 4***). Role-related exits (n = 49) were most commonly attributed to community rejection (n = 23), excessive workload (n = 14), and peer rejection (n = 12). Role-unrelated/life-circumstance exits (n = 55) were most commonly attributed to family duties (n = 24), poor personal health (n = 18), and new employment opportunities (n = 13).

**Table 4.** Reasons for CHW exit by Sex (n=104 who exited)

| Reasons for exit | Female<br>n (%) | Male<br>n (%) | Total |
| --- | --- | --- | --- |
| **Role-related (n=49)** |  |  |  |
| Community rejection | 15 (65.2) | 8 (34.8) | 23 |
| Excessive workload | 8 (57) | 6 (43) | 14 |
| Peer rejection | 5 (41.7) | 7 (58.3) | 12 |
| **Role-unrelated/Life-circumstance (n=55)** |  |  |  |
| Poor personal health | 12 (66.6) | 6 (33.3) | 18 |
| Family duties | 16(66.7) | 8 (33.3) | 24 |
| New employment<br>opportunity | 7 (53.8) | 6 (46.2) | 13 |

## Discussion

This study found that 60.8% of the of the 265 CHWs selected and trained in 2004 to 20028 but still resident in the 3 districts were categorized as retained in their role 17-21 years later. Peer support was the strongest independent predictor of retention (adjusted IRR = 2.21), and consistent or intermittent access to reporting tools also increased the likelihood of retention by approximately 39–46%. The most common reasons cited for exit were family duties, poor personal health, and community rejection.

These findings extend a substantial body of evidence generated within the same Healthy Child Uganda programme. An early impact evaluation of this identical pilot cohort reported favorable retention over its initial years of operation(7), and a subsequent analysis of the same cohort found markedly high medium-term retention of 91% at 2 years and 86% at 5 years (8). Our finding that just over six in ten of the original cohort remained active after 17–21 years indicates a gradual, though far from complete, decline in engagement over an unusually long follow-up period, and situates this cohort among the more durable volunteer CHW programmes reported in the literature, even decades after initial training.

Our identification of peer support as the leading modifiable predictor of long-term retention corroborates earlier qualitative work within the same programme, which linked supportive supervision and constructive peer relationships to sustained CHW performance(10), and is consistent with the broader HCU-derived finding that non-financial incentives, rather than monetary compensation, are the primary drivers of volunteer motivation and continuity(8). A larger, scaled-up HCU cohort similarly reported high medium-term retention driven largely by non-role-related factors rather than financial dissatisfaction (9), although that analysis examined demographic correlates such as sex and age rather than peer support directly. Our results suggest that peer support remains salient even over a much longer follow-up period, during which the influence of early demographic characteristics on retention would ordinarily be expected to decline.

Access to reporting tools has received comparatively little attention in the CHW retention literature, though it aligns with a broader recognition that contextual and health-system-level support shape CHW performance and sustainability(13,14). We interpret consistent or intermittent supply of these simple materials as a proxy for institutional recognition and reduced role burden, rather than as a logistical convenience alone; CHWs who received them may have felt more supported and connected to the formal health system, reinforcing their motivation to continue in the role.

The most common reasons for exit in our cohort - family duties, poor personal health and community rejection - broadly mirror the predominantly ‘logistical’ (role-unrelated) attrition reported in the larger HCU cohort, where relocation, new employment and health problems accounted for the majority of exits (9).

Our findings reinforce that CHW attrition/exit is more often linked to life-course and contextual circumstances than to dissatisfaction with the volunteer role itself (5,6), and that low-cost programmatic investments-fostering peer networks and ensuring a reliable supply of basic reporting materials - may yield durable returns for volunteer CHW programme sustainability over the very long term(14).

### Strengths and limitations

This study’s principal strength is its unusually long follow-up period (17–21 years), which is, to our knowledge, among the longest reported for a volunteer CHW cohort in sub-Saharan Africa, extending prior medium-term evaluations of the same cohort (7,8). The census approach, structured questionnaire, and use of robust variance estimation strengthen the internal validity of our findings.

Nonetheless, several limitations should be considered. First, only CHWs who remained resident in the study districts (265 of the original 404) could be included; those who relocated - a documented common reason for CHW exit in this setting(9) - were necessarily excluded, which may lead to an underestimate of overall attrition or introduce selection bias if migration is itself associated with the exposures under study. Second, our binary definition of retention (regular reporting in the two months preceding data collection) does not capture the quality, intensity, or consistency of CHW engagement over the full 17–21-year period, nor periods of temporary inactivity followed by re-engagement. Third, reasons for exit were self- or peer-reported at a single point in time, in some cases up to two decades after they occurred, and may be subject to recall bias. Finally, as with any single-programme case study, generalizability to other CHW cadres, incentive structures, or health system contexts should be made cautiously.

## Conclusions

More than six in ten volunteer CHWs trained by Healthy Child Uganda between 2004 and 2008 remained actively engaged nearly two decades later, demonstrating that sustained, long-term volunteer CHW engagement is achievable in a low-resource, rural African setting. Peer support and reliable access to basic reporting tools - both low-cost, modifiable programmatic elements - were the strongest independent predictors of this long-term retention. Programme managers and policymakers seeking to sustain CHW cadres over the long term should prioritise structured peer-support mechanisms and consistent provision of essential reporting materials as practical, low-cost strategies for improving the durability of volunteer community health workforces in similar settings.

## Data Availability

All data produced in the present work are contained in the manuscript

## Acknowledgments

We thank the community health workers, district health teams, and community leaders in Mbarara, Rubirizi and Rwampara districts who participated in this study, and the Healthy Child Uganda partnership for its financial support, foundational role in establishing and documenting this cohort.

## HUMAN PARTICIPANTS RESEARCH CHECKLIST

| Item | Details |
| --- | --- |
| **Manuscript title** | Long-term retention and associated factors among volunteer community health workers in southwestern Uganda: a 17-to-21-year retrospective cohort study |
| **Short title** | Long-term CHW retention in Uganda: a 17-to-21-year cohort study |
| **Corresponding author** | Edison Byamugisha |
| **Study design** | Quantitative retrospective cohort study using a census approach; primary data collected by interviewer-administered questionnaire (STROBE) |
| **Study setting** | Mbarara, Rubirizi and Rwampara districts, southwestern Uganda |
| **Data collection** | 1<sup>st</sup> April 2024 to 30<sup>th</sup> April 2024 |

### Summary of Response

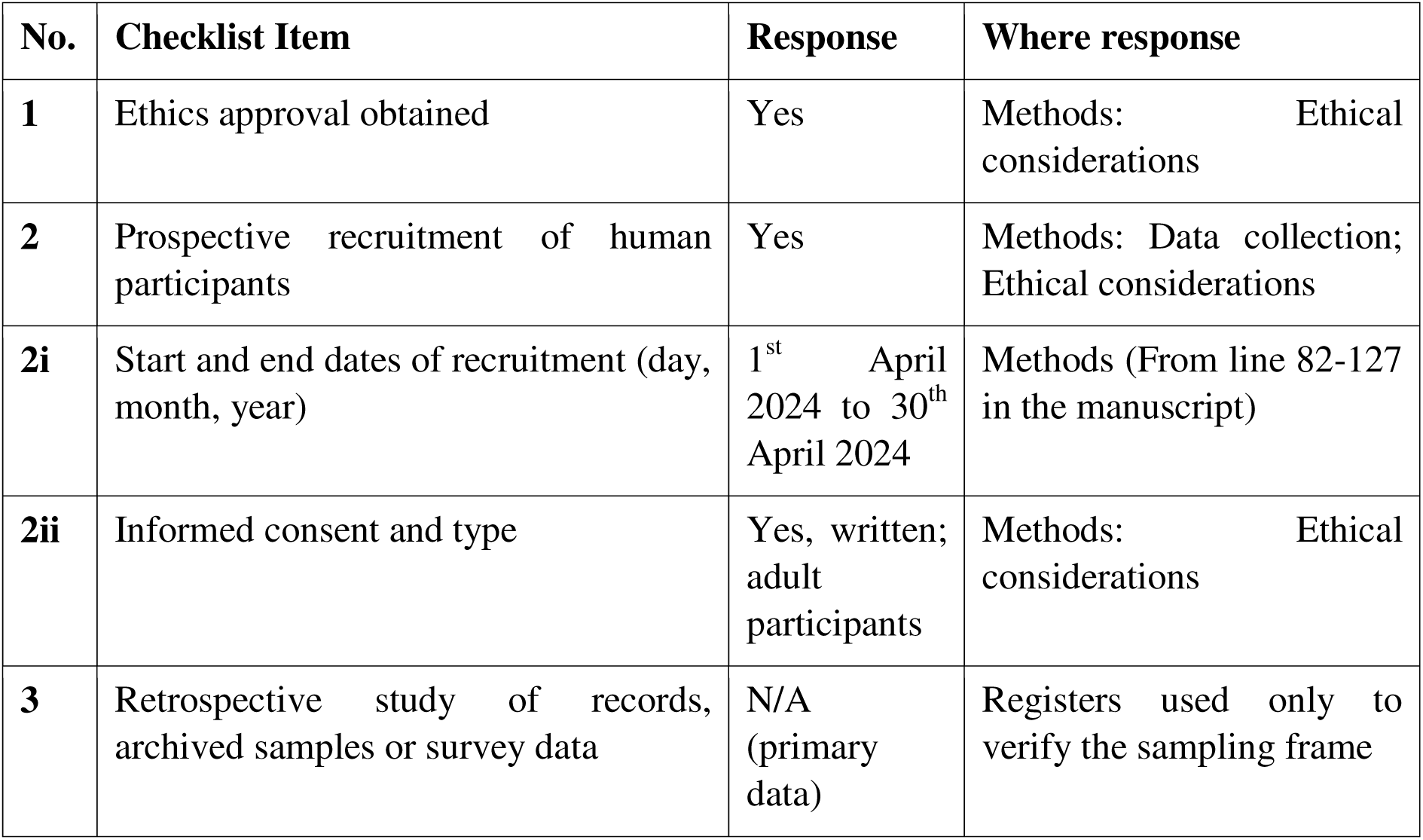

| No. | Checklist Item | Response | Where response |
| --- | --- | --- | --- |
| **1** | Ethics approval obtained | Yes | Methods: Ethical considerations |
| **2** | Prospective recruitment of human participants | Yes | Methods: Data collection; Ethical considerations |
| **2i** | Start and end dates of recruitment (day, month, year) | 1<sup>st</sup> April 2024 to 30<sup>th</sup> April 2024 | Methods (From line 82-127 in the manuscript) |
| **2ii** | Informed consent and type | Yes, written; adult participants | Methods: Ethical considerations |
| **3** | Retrospective study of records, archived samples or survey data | N/A (primary data) | Registers used only to verify the sampling frame |

### 1. Ethical Approval

**Did you obtain ethics approval for this study?** Yes.

Ethical approval was obtained from the following bodies, as reported in the Methods (Ethical considerations):

| Approving body | Reference Number | Document to upload |
| --- | --- | --- |
| Research Ethics Committee, Mbarara University of Science and Technology | MUST-2023-1183 | Original approval letter (file type “Other”) |
| Uganda National Council for Science and Technology | SS2542ES | Original approval letter (file type “Other”) |
| Administrative clearance | From three districts – Mbarara, Rubirizi and Rwampara Districts | Original administrative clearance(file type “other”) |
| Informed consent forms | All eligible participants | Original informed consent form(file type”other”) |

- Upload the original approval documents, not any extension or renewal letters.
- If either document is not in English, provide an English translation.
- Data collection took place only in Uganda; therefore the explanation required for international studies without approval from all countries does not apply.
- Community entry approvals were also obtained from the Chief Administrative Officers of the three districts. These are not ethics approvals but may be mentioned for completeness.

**Explanation for absence of ethics approval:** Not applicable.

### 2. Prospective Recruitment of Participant

**Did you prospectively recruit human participants?** Yes. Community health workers (CHWs) were recruited directly and interviewed in 1^st^ April 2024 to 30^th^ April 2024 using a pretested, structured, interviewer-administered questionnaire. Although the design is described as a retrospective cohort, the participants were recruited and interviewed specifically for this study.

| Requirement | Response |
| --- | --- |
| **i. Recruitment period** | 1<sup>st</sup> April 2024 to 30<sup>th</sup> April 2024 |
| **ii. Informed consent** | Yes. Participants provided written informed consent before enrolment. A copy of the original consent form has been attached. |
| **Minors** | Not applicable. All participants were adults (median age 55 years, IQR 50–60), so parental or guardian consent was not required. |
| **Confidentiality** | Each participant was assigned a unique code and names were not recorded. Participants received a transport reimbursement of UGX 10,000 (USD 2.70). |
| **Line numbers in Methods** | From line 82-127 in the manuscript. |

### 3. Retrospective Study of Records, Archived samples or Survey Data

**Is this a retrospective study of existing records or archived data?** Not applicable to the main study. All study data were collected directly from participants during 1^st^ April 2024 to 30^th^ April 2024.

| **Requirement** | **Response** |
| --- | --- |
| **ii. Date data were accessed** | Not applicable for the study data. CHW registers maintained since 2004 were used by parish coordinators and investigators only to verify and update the list of eligible CHWs (sampling frame). |
| **iii. Access to identifying information** | Participant names were not recorded and each participant was assigned a unique code. Only the principal investigator had access to names and contacts during data collection and afterwards. |
| **Line numbers in Methods** | From line 82-127 in the manuscript. |

### STROBE Statement-Checklist of Items to be included in Reports of Cohort Studies

*Manuscript: “Long-term retention and associated factors among volunteer community health workers in southwestern Uganda: a 17-to-21-year retrospective cohort study”* Completed by: Edison Byamugisha, Corresponding author

| Item No | Recommendation | Reported in Manuscript (Section /location) |
| --- | --- | --- |
| **Title and Abstract** |  |  |
| 1a | Indicate the study's design with a commonly used term in the title or the abstract | Title (“...a 17-to-21-year retrospective cohort study”); Abstract, sentence 3 |
| 1b | Provide in the abstract an informative and balanced summary of what was done and what was found | Abstract |
| **Introduction** |  |  |
| 2 | Explain the scientific background and rationale for the investigation being reported | Introduction, paragraphs 1–3 |
| 3 | State specific objectives, including any prespecified hypotheses | Introduction, final paragraph |
| **Methods** |  |  |
| 4 | Present key elements of study design early in the paper | Methods, “Study design and setting” |
| 5 | Describe the setting, locations, and relevant dates, including periods of recruitment, exposure, follow-up, and data collection | Methods, “Study design and setting” (districts, 2004–2008 training period); “Data collection” (April 2024) |
| 6a | Give the eligibility criteria, and the sources and methods of selection of participants. Describe methods of follow-up | Methods, “Study population and eligibility” |
| 6b | For matched studies, give matching criteria and number of exposed and unexposed | Not applicable – not a matched study |
| 7 | Clearly define all outcomes, exposures, predictors, potential confounders, and effect modifiers. Give diagnostic criteria, if applicable | Methods, “Outcome definition” (retention); predictor variables (peer support, reporting-tool access, sex, age) introduced in Results, Table 2, but not separately defined as a priori |
|  |  | exposures in Methods |
| 8 | For each variable of interest, give sources of data and details of methods of assessment (measurement). Describe comparability of assessment methods if there is more than one group | Methods, “Data collection” (pretested, structured, interviewer-administered questionnaire, translated into Runyankole; single interviewer, EB) |
| 9 | Describe any efforts to address potential sources of bias | Methods, “Study population and eligibility” (census approach, avoiding sampling bias) and “Data collection” (pretested, standardized, single-interviewer instrument); residual bias discussed in Discussion, “Strengths and limitations” |
| 10 | Explain how the study size was arrived at | Methods, “Study population and eligibility” (census of entire eligible cohort, n = 404, chosen because the population was manageable) |
| 11 | Explain how quantitative variables were handled in the analyses. If applicable, describe which groupings were chosen and why | Partially reported – age reported as median with IQR in Results, Table 2; not categorized into groups, and no explicit rationale given for treating it continuously |
| 12a | Describe all statistical methods, including those used to control for confounding | Methods, “Statistical analysis” (bivariate and multivariable Poisson regression with robust standard errors; purposeful selection per Hosmer and Lemeshow) |
| 12b | Describe any methods used to examine subgroups and interactions | Methods, “Statistical analysis” (interaction term between community type and district tested by log- |
|  |  | likelihood ratio test, found non-significant and dropped) |
| 12c | Explain how missing data were addressed | Not explicitly reported |
| 12d | If applicable, explain how loss to follow-up was addressed | Not addressed as a statistical method; the 139 CHWs who had relocated and could not be followed up are discussed narratively in Discussion, “Strengths and limitations” |
| 12e | Describe any sensitivity analyses | Not reported |
| **Results** |  |  |
| 13a | Report numbers of individuals at each stage of study | Results, “Cohort disposition” and Table 1 (404 originally trained - 265 still resident - 161 retained / 104 exited) |
| 13b | Give reasons for non-participation at each stage | Not reported – reasons are given for exit from the programme (Table 4), but not for any non-participation/non-consent among the 265 resident CHWs approached |
| 13c | Consider use of a flow diagram | Not included; cohort flow is presented narratively and in Table 1 only |
| 14a | Give characteristics of study participants and information on exposures and potential confounders | Results, “Characteristics of the participants” and Table 2 |
| 14b | Indicate number of participants with missing data for each variable of interest | Not reported |
| 14c | Summarise follow-up time (e.g., average and total amount) | Follow-up expressed as the fixed interval since training (17–21 years; title, Abstract, Introduction); no mean/median follow-up time per participant is |
|  |  | reported in Results |
| 15 | Report numbers of outcome events or summary measures over time | Results, “Cohort disposition” and Table 1 (161/265, 60.8% retained) |
| 16a | Give unadjusted estimates and, if applicable, confounder-adjusted estimates and their precision. Make clear which confounders were adjusted for and why | Results, Table 2 (unadjusted IRR) and Table 3 (adjusted IRR, 95% CI); text under “Multivariable analysis of factors associated with retention”; Table 3 footnote notes adjustment for sex and age |
| 16b | Report category boundaries when continuous variables were categorized | Not applicable – age was analysed as a continuous variable and not categorized |
| 16c | If relevant, consider translating estimates of relative risk into absolute risk for a meaningful time period | Not reported |
| 17 | Report other analyses done – e.g. analyses of subgroups and interactions, and sensitivity analyses | Results, “Reasons for exit” and Table 4 (exit reasons stratified by sex); Methods, “Statistical analysis” (community type × district interaction test, reported as non-significant) |
| **Discussion** |  |  |
| 18 | Summarise key results with reference to study objectives | Discussion, paragraph 1 |
| 19 | Discuss limitations of the study, taking into account sources of potential bias or imprecision. Discuss both direction and magnitude of any potential bias | Discussion, “Strengths and limitations” |
| 20 | Give a cautious overall interpretation of results considering objectives, limitations, multiplicity of analyses, results from similar studies, and other relevant evidence | Discussion, paragraphs 2–5 (comparison with earlier HCU cohort evaluations and wider CHW literature) |
| 21 | Discuss the generalizability (external validity) of the study results | Discussion, “Strengths and limitations”, final sentence (generalizability to other CHW cadres/contexts should be |
|  |  | made cautiously) |
| **Other information** |  |  |
| 22 | Give the source of funding and the role of the funders for the present study and, if applicable, for the original study on which the present article is based | Acknowledgments (Healthy Child Uganda partnership acknowledged for financial support and its foundational role in establishing the cohort); specific role of the funder in the design, conduct, analysis or reporting of the present study is not separately detailed |

\*Give information separately for exposed and unexposed groups.
**Note:** An Explanation and Elaboration article discusses each checklist item and gives methodological background and published examples of transparent reporting. The STROBE checklist is best used in conjunction with this article (freely available on the Web sites of PLoS Medicine at http://www.plosmedicine.org/, Annals of Internal Medicine at http://www.annals.org/, and Epidemiology at http://www.epidem.com/). Information on the STROBE Initiative is available at http://www.strobe-statement.org.

---

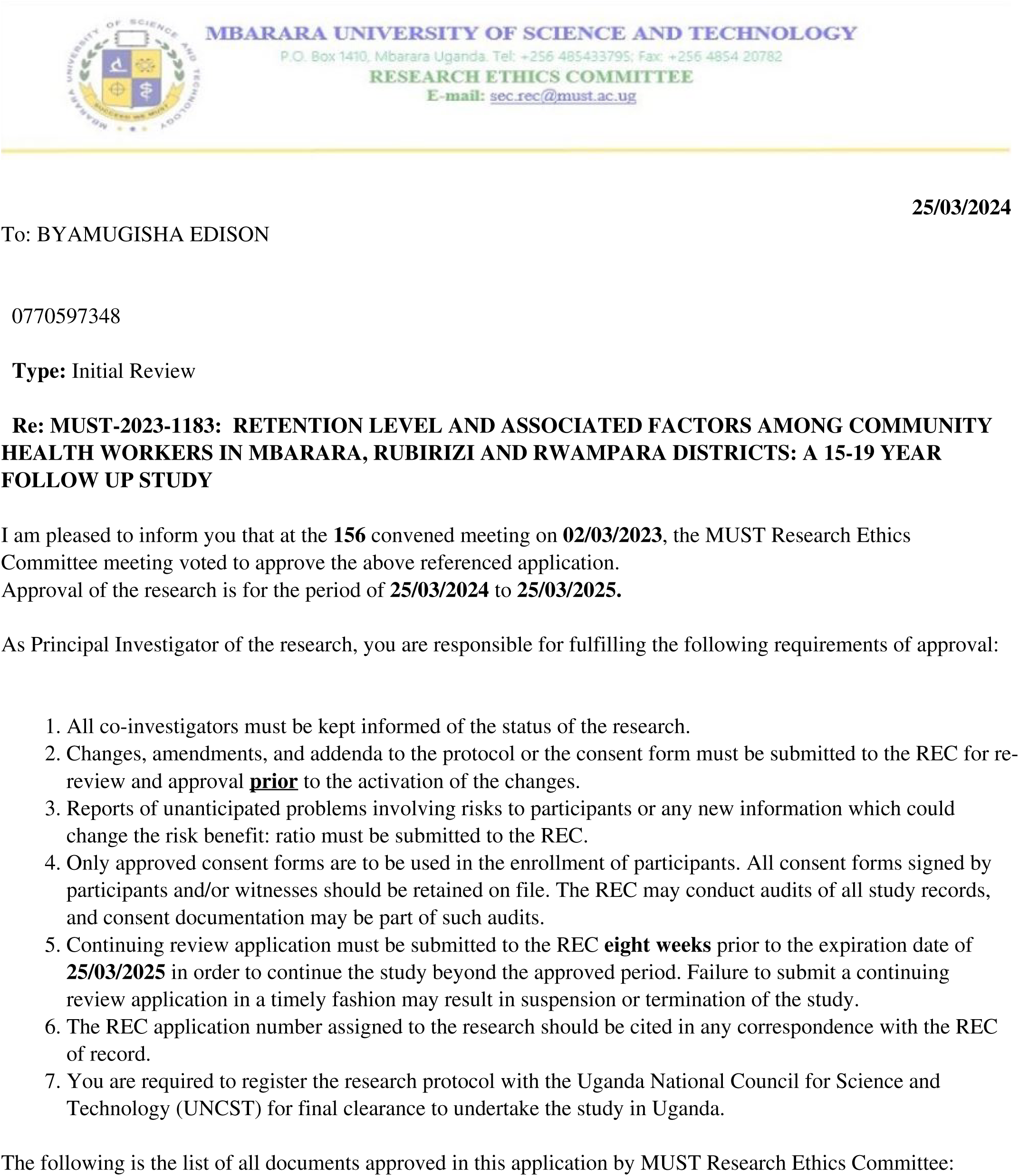

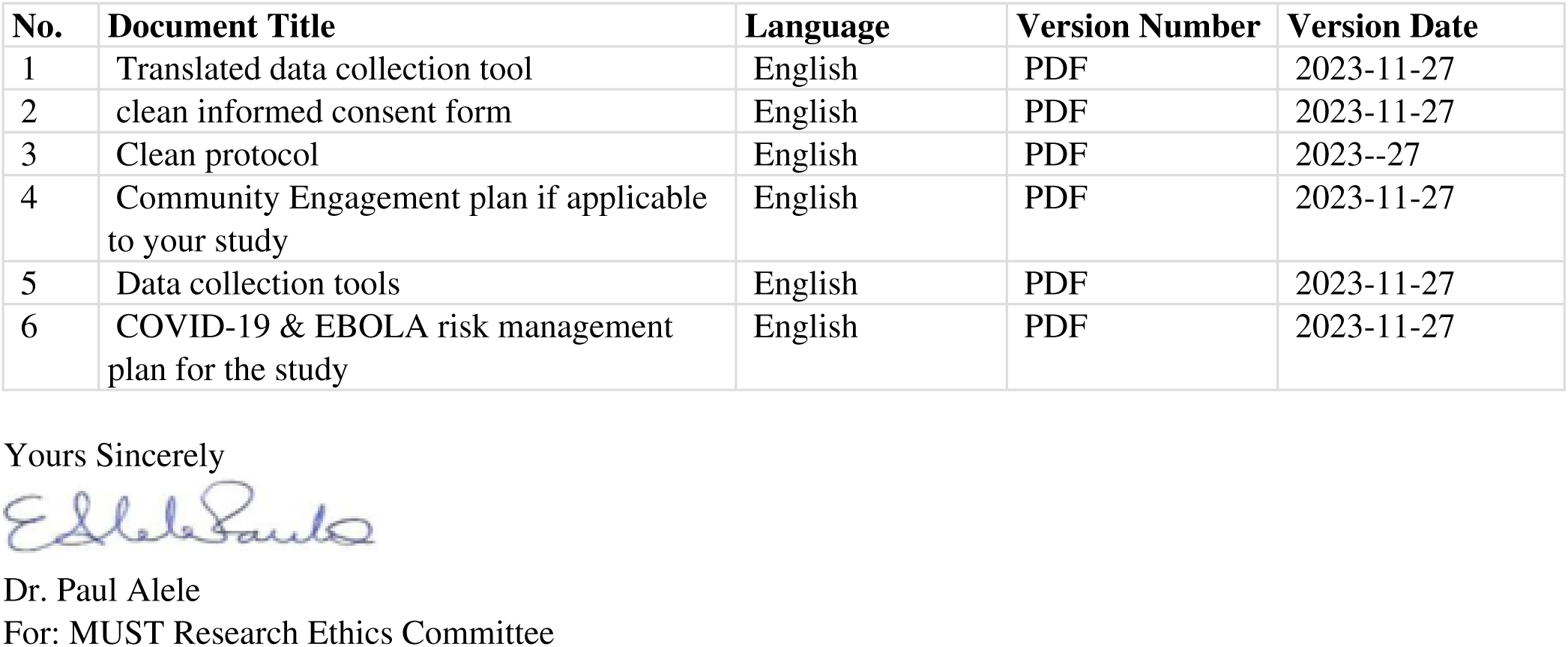

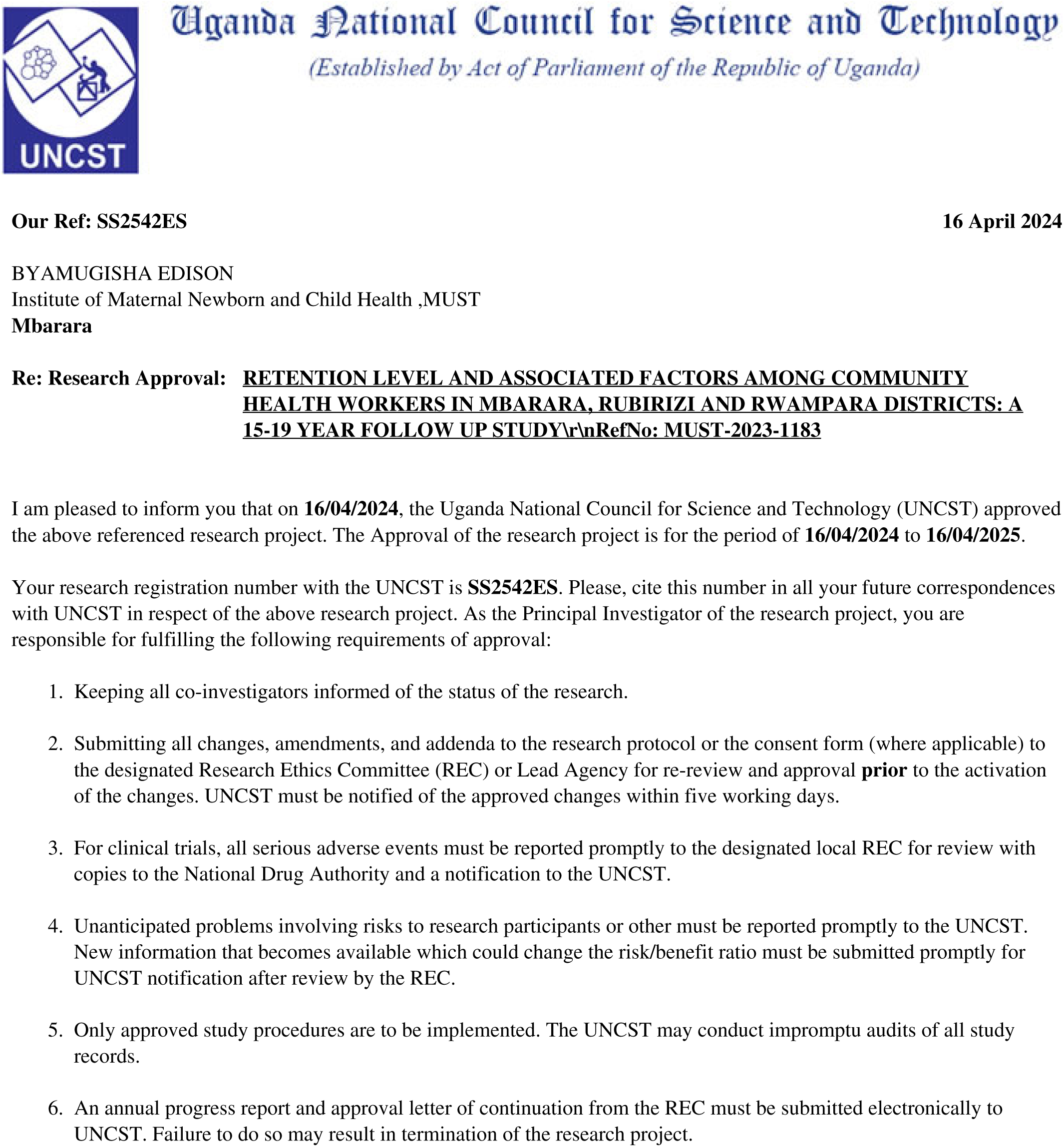

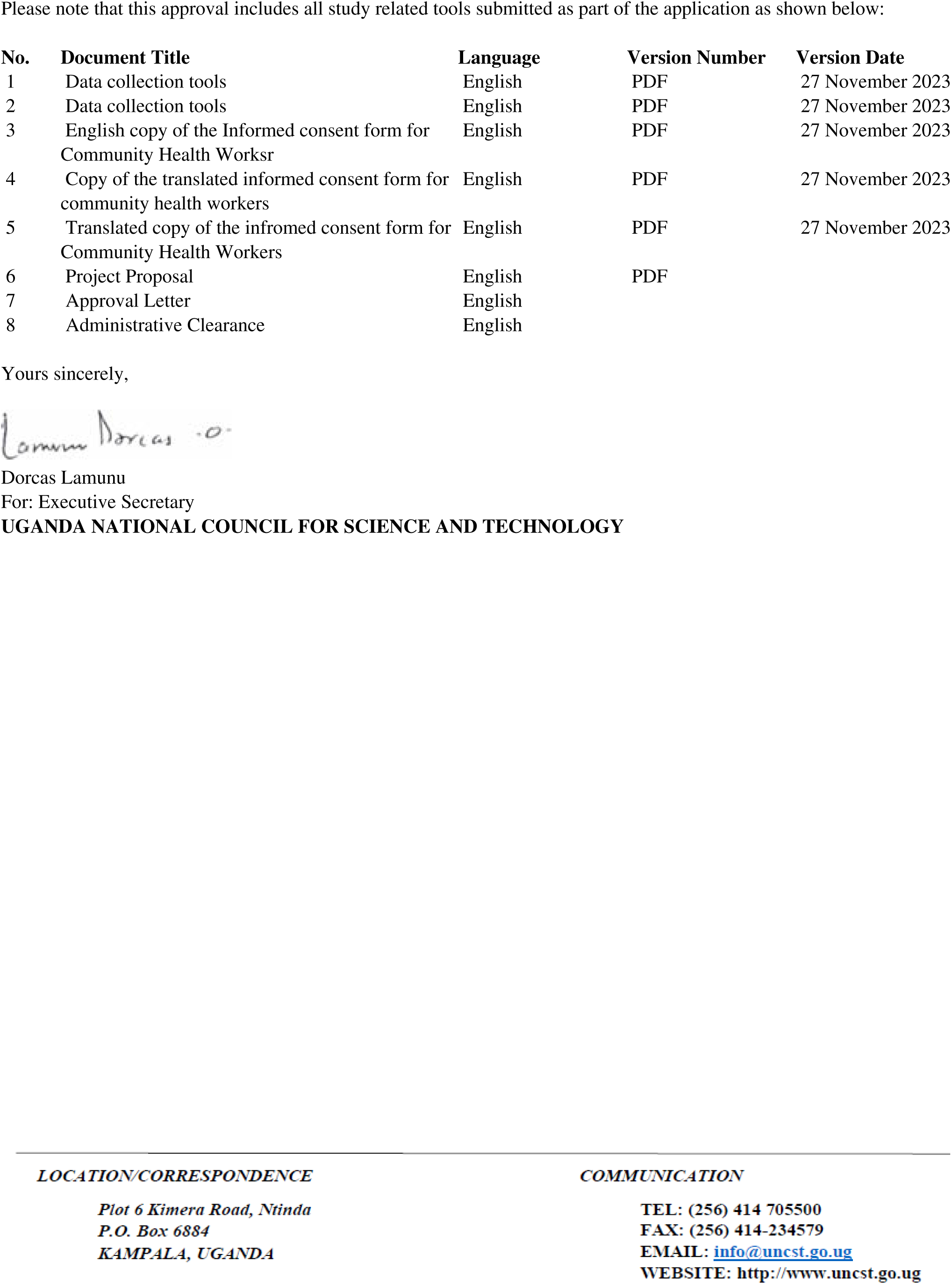

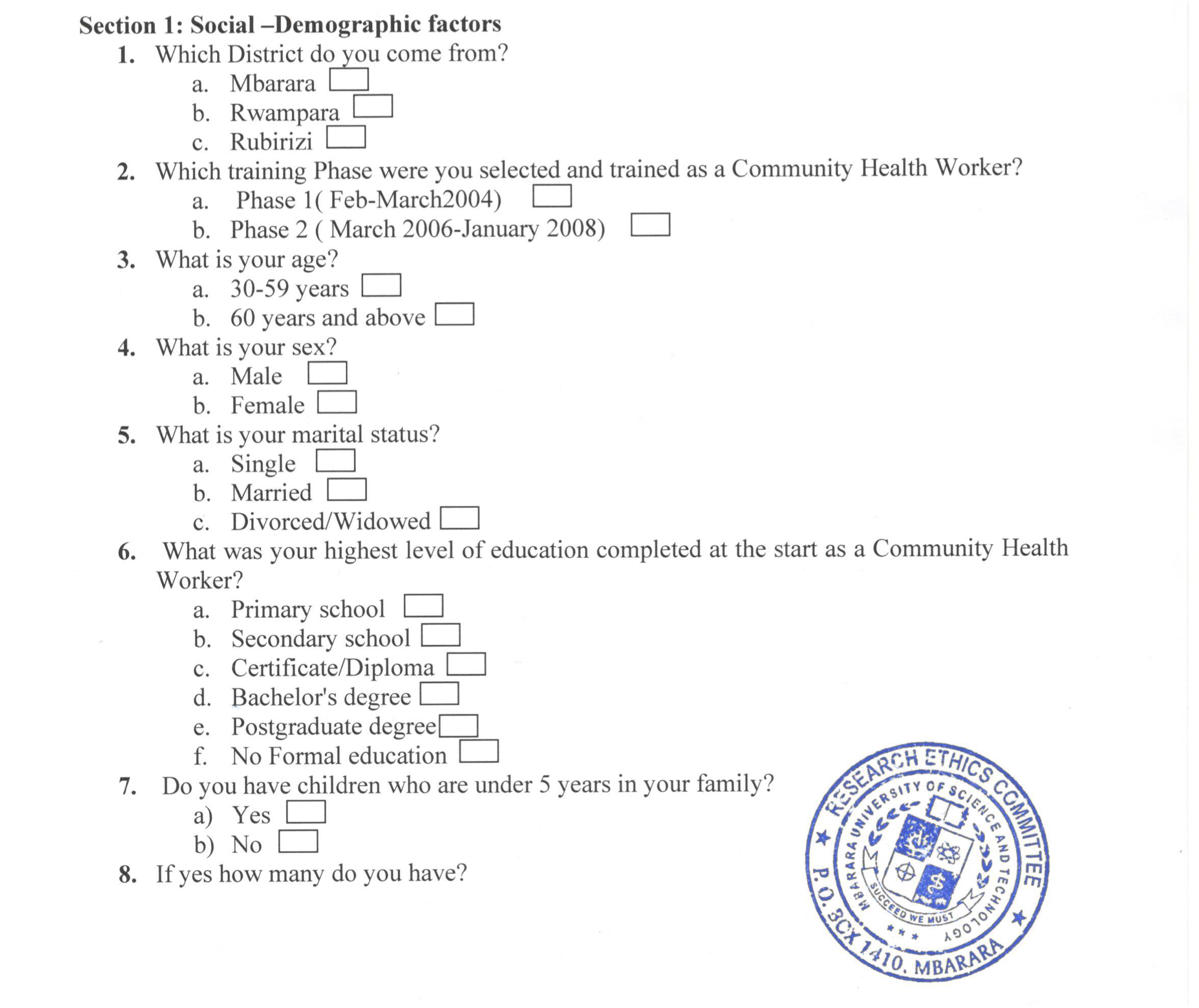

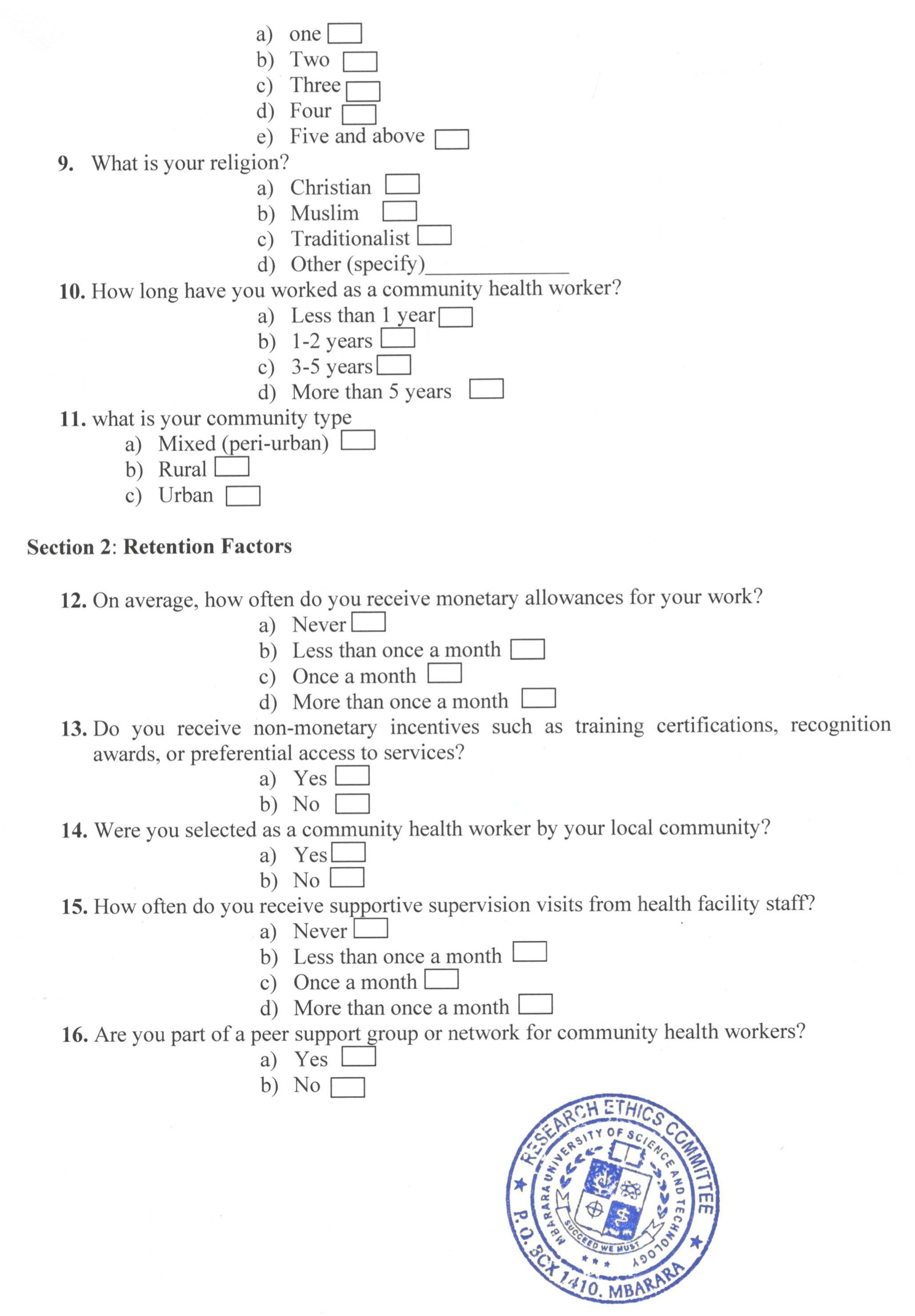

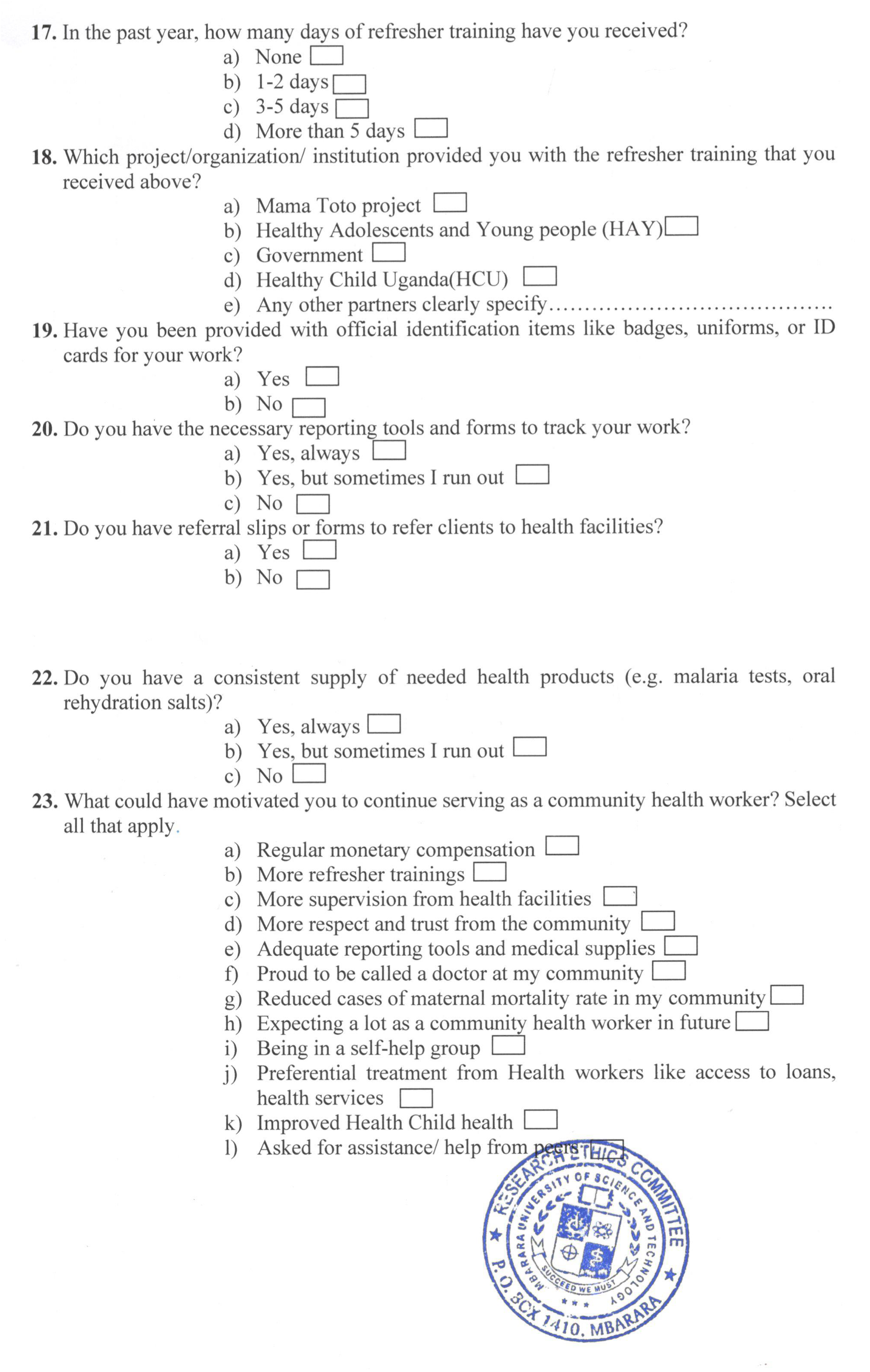

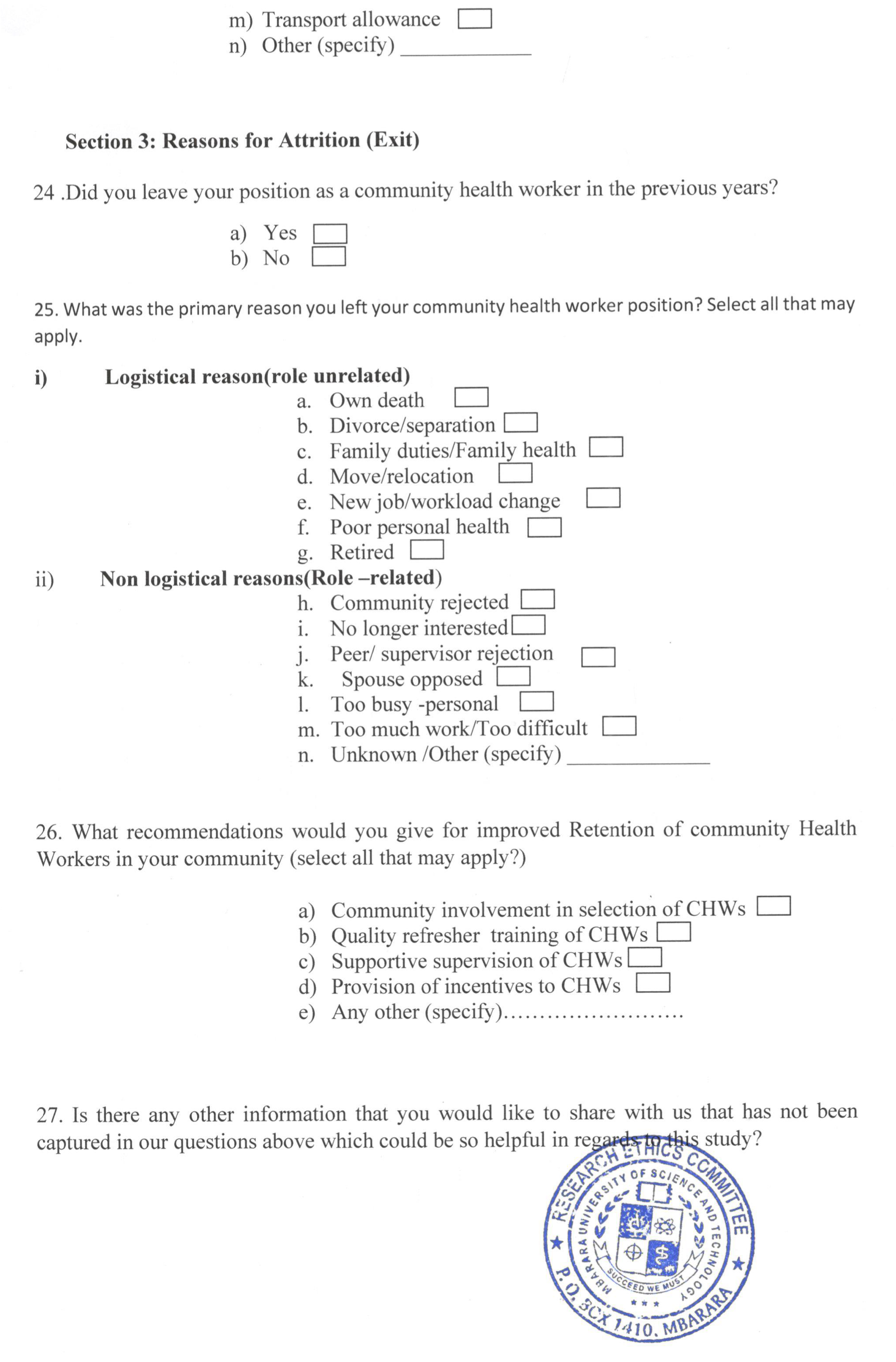

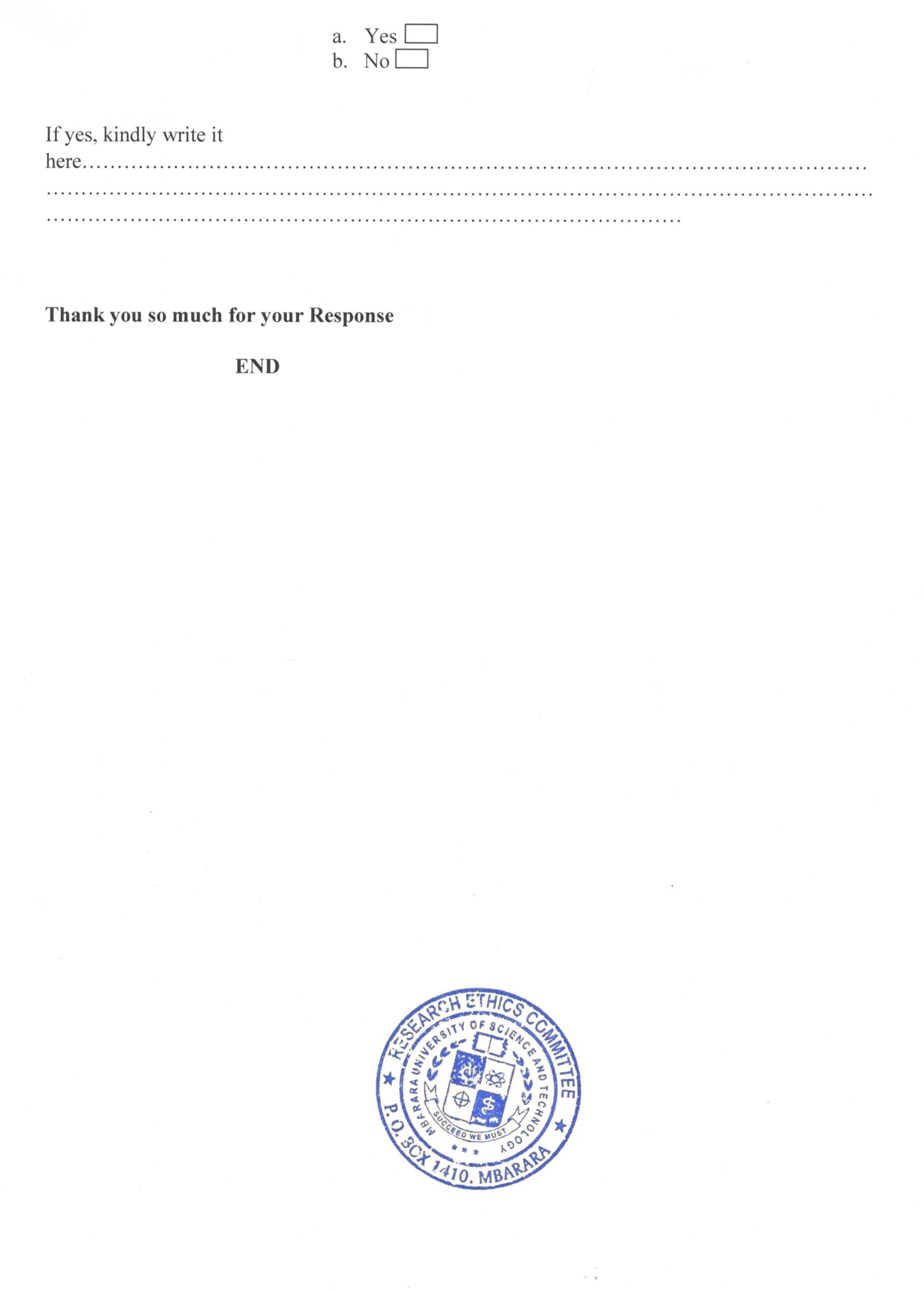

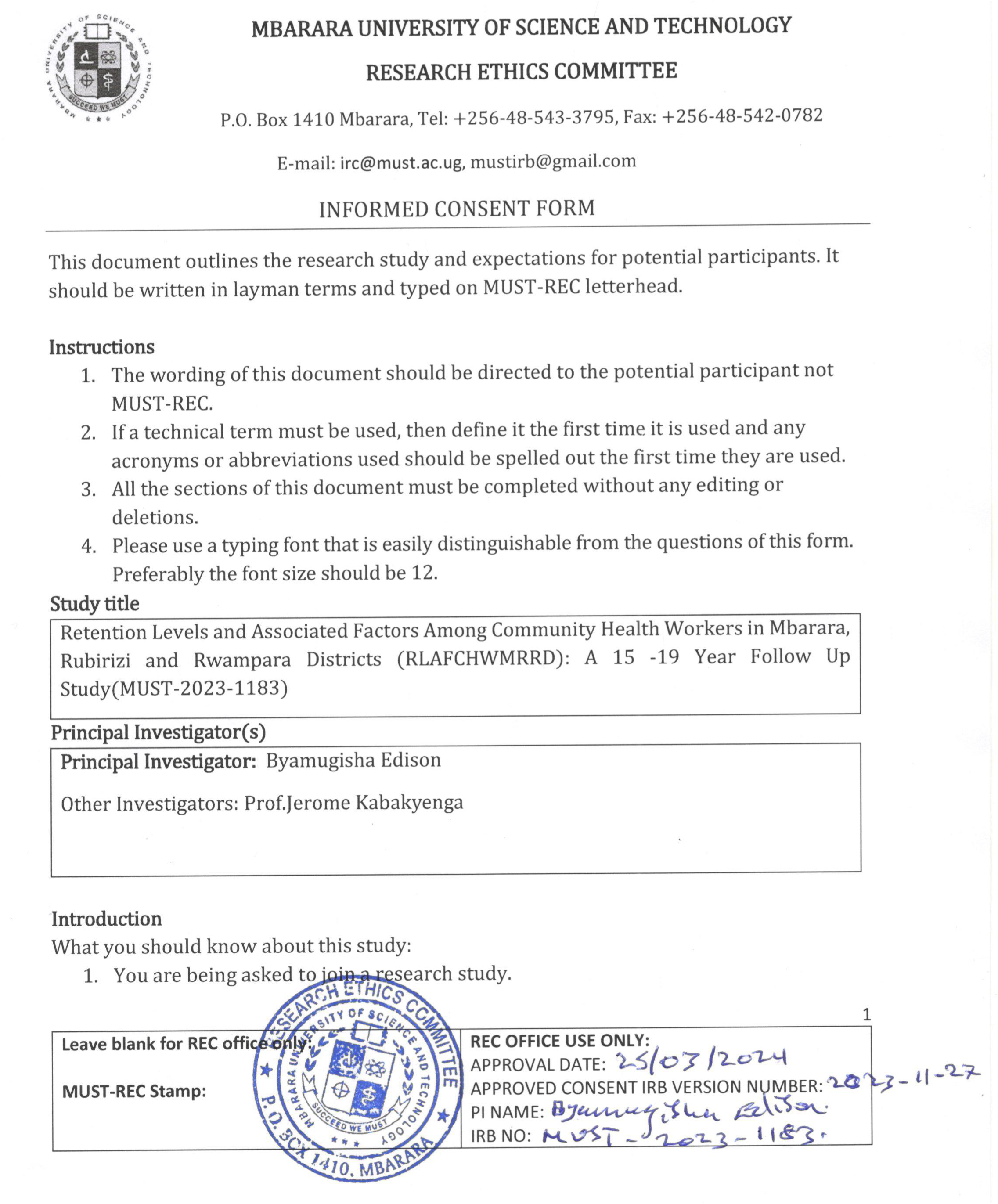

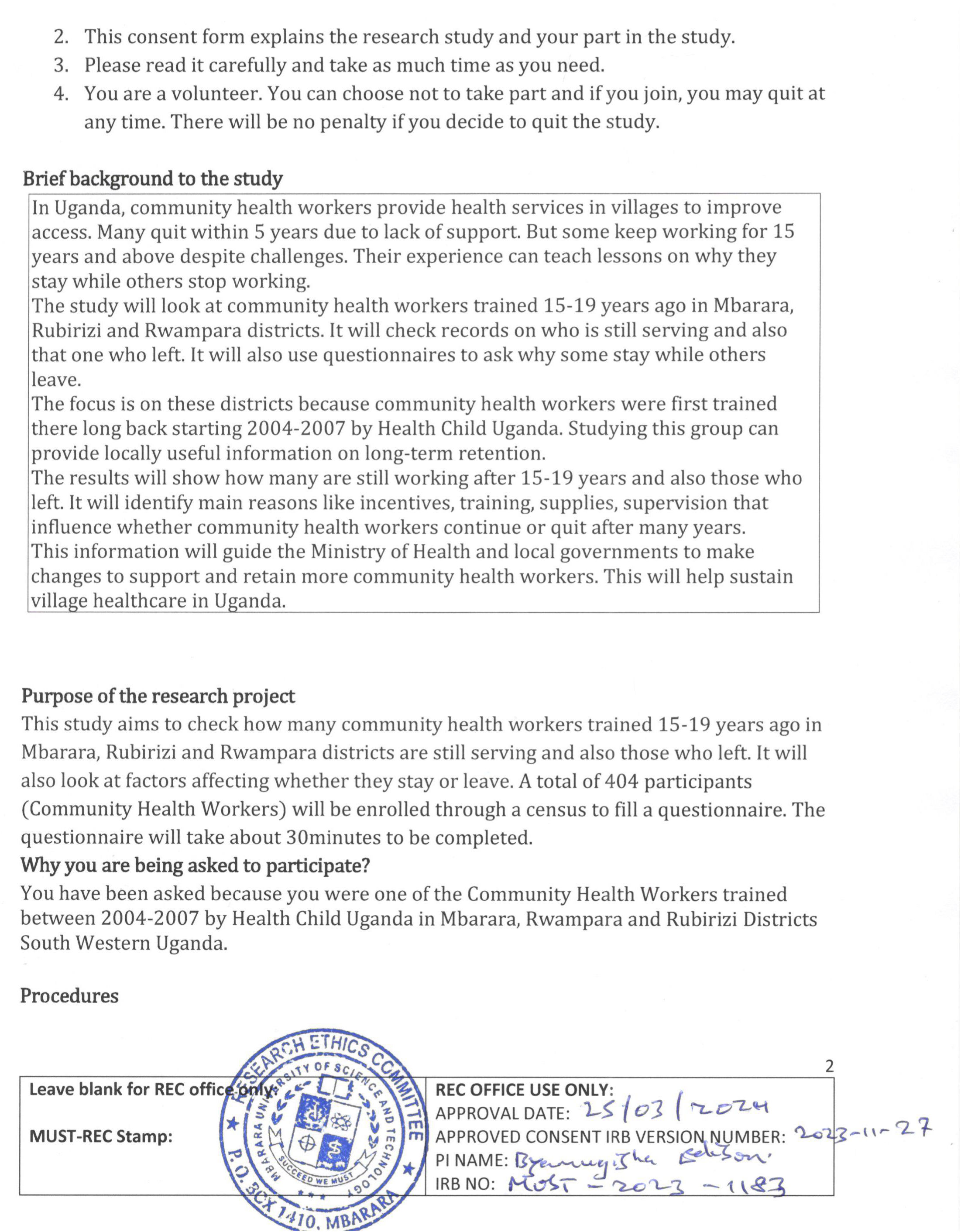

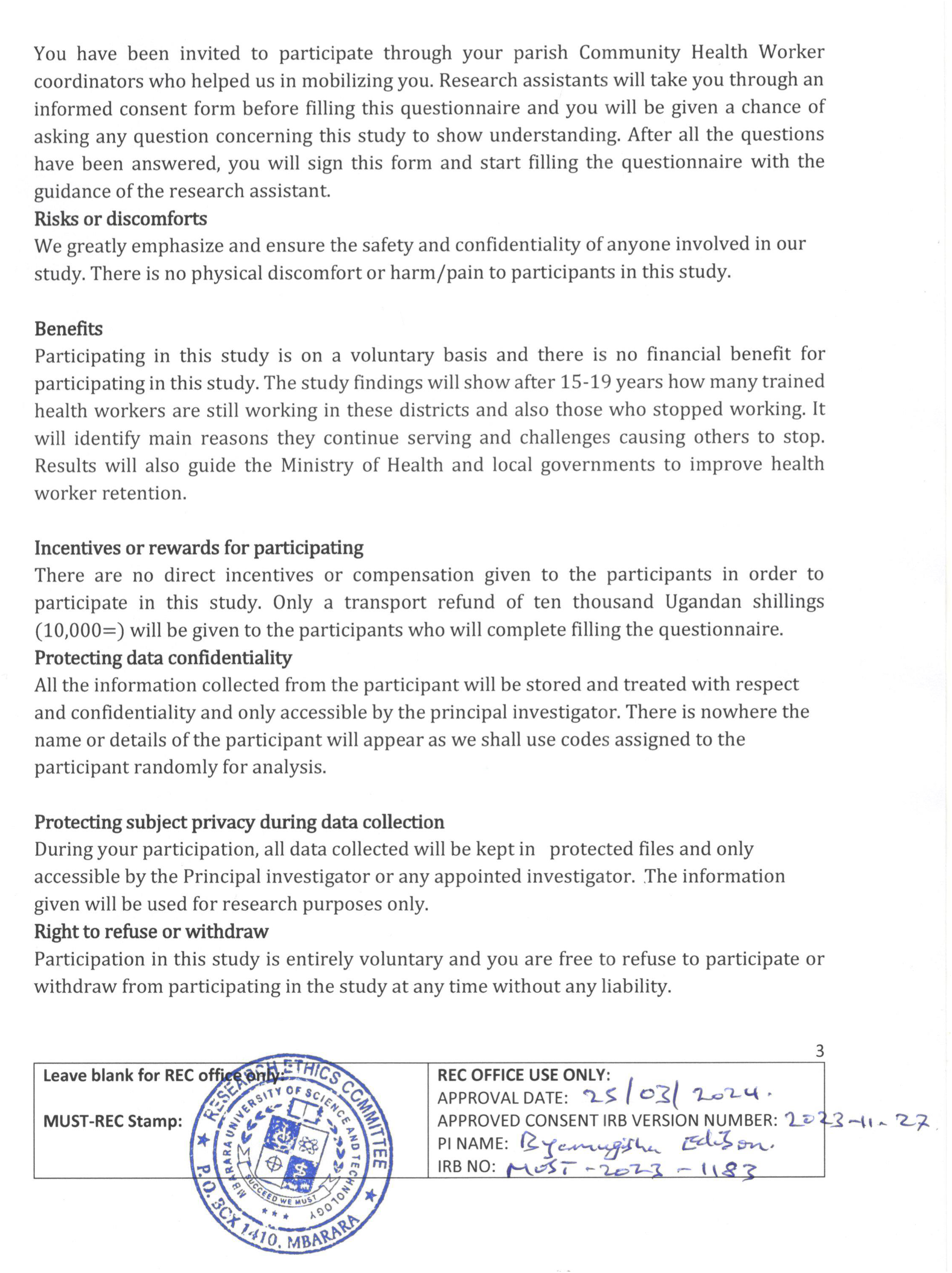

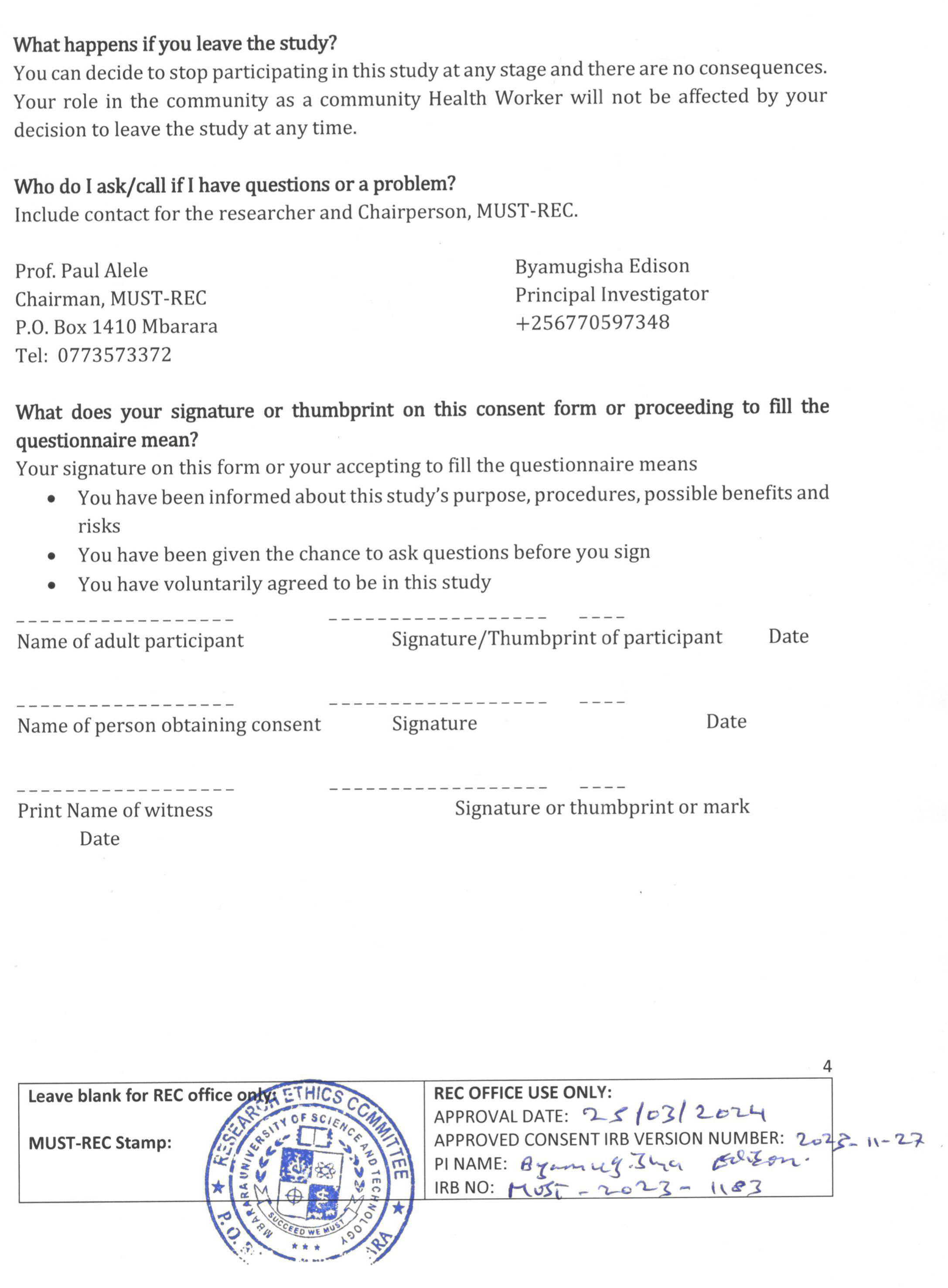

